# Transcranial Light Stimulation Improves Selective Attention in Children with ADHD

**DOI:** 10.1101/2025.09.18.25335086

**Authors:** Yongheng Zhao, Yang Li, Keyao Zhang, Zhilin Li, Yiqing Hu, Lirou Tan, Hai Jia, Shaodi Wang, Zhibing Gao, Yan Song, Xiaoli Li, Chenguang Zhao, Aihua Cao

**Affiliations:** Department of Pediatrics, Qilu Hospital of Shandong University, Jinan, 250012, Shandong Province, China; Beijing Institute for Brain Research, Chinese Academy of Medical Sciences & Peking Union Medical College, Beijing, 102206, China; Chinese Institute for Brain Research, Beijing, Beijing, 102206, China; School of Biomedical Engineering, Faculty of Medicine, Dalian University of Technology, Dalian, 116024, China; Biomedical Engineering Department, University of Texas at Arlington, Arlington, 76019, USA; Department of Anesthesiology, Peking University Third Hospital, Beijing, 100191, China; National Center for Mental Health, Beijing, China; State Key Laboratory of Cognitive Neuroscience and Learning, Beijing Normal University, Beijing, 100875, China

**Author notes:** These authors contributed equally to this work (Yongheng Zhao, Yang Li, Keyao Zhang).

**Keywords:** transcranial Light Stimulation (tLS), Children with Attention Deficit Hyperactivity Disorder (ADHD), Selective Attention, Electroencephalography (EEG), transcranial photobiomodulation (tPBM)

## Abstract

Children with attention deficit/hyperactivity disorder (ADHD) frequently display impairments in selective attention. Prior research has identified the dorsolateral prefrontal cortex (dlPFC) as a critical region contributing to these deficits. In this study, we examined the effects of transcranial light stimulation (tLS) over the right dlPFC on selective attention in children with ADHD. In Experiment 1, we modeled photon propagation pathways and attenuation patterns within pediatric cranial structures to determine the optimal stimulation dose, estimating that the effective tLS dosage in children should be approximately 40% of the adult level. In Experiment 2, we applied these parameters in a triple-blind, randomized, crossover clinical trial involving 40 children with ADHD. Active tLS significantly increased the amplitude of the event-related potential (ERP) markers of selective attention and strengthened spatial Selective cortical tuning function (CTF) modulation. These neural changes predicted measurable improvements in attention symptoms over the subsequent following week. In Experiment 3, we further included typically developing (TD) children as a control group and found that the selective attention performance in children with ADHD following active tLS approached the levels observed in TD children. Our findings demonstrate that right dlPFC-targeted tLS enhances selective attention in children with ADHD and produces clinically meaningful improvements in inattention. This study provides novel evidence supporting the potential of tLS as a therapeutic intervention for ADHD.

## Introduction

Attention-deficit/hyperactivity disorder (ADHD) is a prevalent neurodevelopmental condition in children, with an estimated global prevalence of approximately 5.3% ^1,2^. It is characterized by the core symptoms of inattention, hyperactivity, and impulsivity^3^. ADHD often persists into adolescence and adulthood, underscoring the importance of early intervention in pediatric populations. Given the high neuroplasticity of the developing brain, timely treatment may more effectively alleviate symptoms and reduce long-term adverse outcomes^2^. Psychostimulant medications remain the standard first-line treatment; however, they present notable limitations, including variable efficacy, safety concerns, and challenges with long-term adherence. Many children experience adverse side effects or insufficient symptom control ^4^, leading to parental reluctance to pursue medication. These challenges highlight the urgent need for safe and effective non-pharmacological interventions for ADHD. Non-invasive brain stimulation (NIBS) approaches, such as repetitive transcranial magnetic stimulation (rTMS) and transcranial direct current stimulation (tDCS), have shown preliminary promise in modulating dysfunctional neural circuits, The current-based ^5,6^ and magnetic field-based stimulation ^7,8^ of the dorsolateral prefrontal cortex (dlPFC) remains the most extensively studied target for improving attention. While some studies report beneficial effects on specific cognitive functions, current evidence ^9^ remains limited and insufficient to establish these techniques as clinically viable alternatives.

Transcranial light stimulation (tLS) has emerged as a promising non-pharmacological intervention with potential clinical applications. Preclinical animal studies have shown that tLS enhances oxidative phosphorylation efficiency and increases adenosine triphosphate (ATP) synthesis. At the cellular level, cytochrome c oxidase (CCO) - the terminal enzyme in the mitochondrial electron transport chain - serves as the primary photon acceptor for this photobiomodulation process. Recent clinical investigations further support the efficacy of tLS in ameliorating symptoms across multiple domains, including cognition, attention, social interaction, and behavior, in conditions such as dementia ^10^, depression ^11^ and autism spectrum disorders ^12–14^. tLS typically employs red to near-infrared wavelengths (600-1100 nm), which can penetrate the scalp and skull of human to target cortical brain regions. This mechanism may also modulate the neurovascular unit ^15^ and enhance cognitive functions ^16^, suggesting therapeutic potential for ADHD, particularly in domains of attention ^17^, working memory ^16^, and emotion regulation ^18^. Despite this promise, two major challenges remain in applying tLS as a non-pharmacological intervention for children with ADHD.

First, a recent behavioral study reported that tLS applied to the dlPFC significantly improved working memory and attention performance in adults with ADHD ^19^. However, it remains unclear whether stimulation parameters optimized for adults can effectively modulate attention in children. Human studies have shown that photon attenuation during tLS varies developmentally, as age-related differences in skin and skull thickness significantly affect light penetration ^20^. While evidence suggests that tLS delivered at approximately 250 mW/cm² can modulate neuronal activity in adults ^21^, the optimal pediatric dosage - one that produces the predicted neural effects while ensuring safety, has yet to be established for children and adolescents. Second, growing evidence indicates that both short- and long-term interventions for ADHD may require diverse measurement approaches to accurately capture treatment effects ^9^. Although rating scales remain the primary clinical assessment tool, their subjective nature compromises reliability and validity ^22^, and they may not adequately detect the subtle neuromodulatory effects of NIBS. This limitation has encouraged the search for more objective measures, with EEG research successfully identifying several neural biomarkers characteristic of ADHD. In particular, children with ADHD show distinct patterns in parieto-occipital N2pc components and alpha-band oscillations. These EEG markers consistently reflect selective attention deficits compared with those of typically developing (TD) peers^23,24^_._

To address these gaps in applying tLS to children with ADHD, the present study comprises three experiments (**Fig. 1A**). In Experiment 1, individualized pediatric head models were used to simulate photon distribution and determine safe, effective dosing parameters tailored for children, as existing stimulation protocols are based on adult anatomy. In Experiment 2, a randomized, triple-blind, sham-controlled design incorporating multimodal assessments (EEG, eye-tracking, cognitive performance, and clinical measures) was employed to clarify the neurophysiological mechanisms of tLS and identify attention-related biomarkers distinguishing active from sham stimulation. In experiment 3, a TD control group was added to directly compare patients with ADHD with and without tLS treatment against age-matched normative baselines. This allowed us to evaluate whether tLS not only improves attention and clinical symptoms in ADHD but also normalizes their cognitive and neural profiles toward those of TD peers. We hypothesized that tLS applied to the right dlPFC would ameliorate deficits in selective attention in children with ADHD, as measured through improvements in both behavioral performance and neural markers. In light of anatomical differences between pediatric and adult populations, we anticipated that a calibrated reduction in stimulation dosage relative to adult standards would be necessary to achieve effective and safe neuromodulation. Furthermore, we expected that active tLS would not only enhance attentional performance compared to sham stimulation but also normalize neural and behavioral profiles in children with ADHD toward levels observed in TD group, thereby supporting the therapeutic potential of tLS as a safe and effective non-pharmacological intervention for ADHD.

**Fig. 1.**
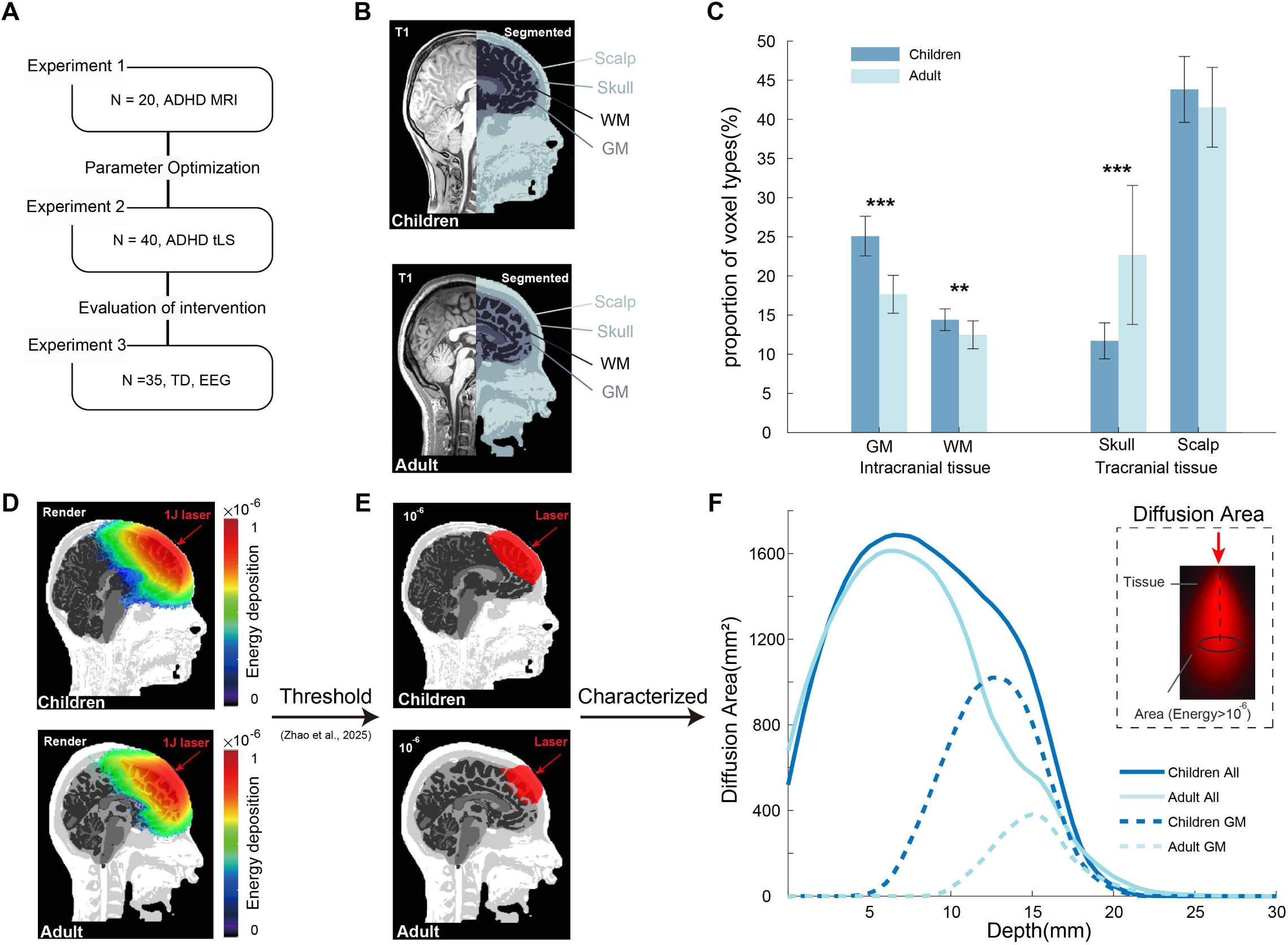
Study framework and comparison of tissue composition and intracranial photon energy distribution between children and adults. A. Study framework illustrating the three main experiments, including subject numbers and primary objectives. B. Representative subjects from the two groups, showing raw T1-weighted images (the left half) and segmentation into multiple tissue types (the right half). C. Group comparison of voxel proportions, with significant differences observed between children and adults in gray matter, white matter, and skull. Statistical significance was assessed using two-tailed independent-samples t-tests (**p<0.01, ***p<0.001). D. Monte Carlo–based intracranial photon energy deposition maps in children and adults under identical incident dose conditions (1 J). E. Distribution of voxels exceeding the activation threshold (10^-6^) under identical incident dose conditions for the two groups. F. Depth-resolved diffusion area (mm²), calculated in each plane orthogonal (upper right) to the incident beam path by summing the voxel area where energy deposition exceeded the predefined threshold. Depth was measured from the center of the projected beam footprint on this scalp surface. upper right: Spatial distribution of intracranial energy deposition across the depth and lateral directions aligned with the incident path.

## Materials and Methods

### Study Design

In experiment 1, we aimed to determine the appropriate power density of tLS for children with ADHD by calculating intracranial photon energy deposition in children compared with adults. A dosimetry study was conducted to simulate the dose translation from adults to children. Tissue segmentation was performed using the CAT12 toolbox, which generated probabilistic maps for six tissue types: air, scalp, skull, cerebrospinal fluid (CSF), gray matter (GM), and white matter (WM). Each voxel was assigned a unique tissue label based on its highest probability, resulting in an individual anatomical map with discrete tissue classification (**Fig. 1B**). To assess group-level differences in tissue composition, voxel proportions of GM, WM, skull, and scalp were calculated for each subject. A two-tailed statistical comparison was then performed between the pediatric and adult groups to evaluate potential differences in intracranial and extracranial compartments (**Fig. 1C**).

Based on the individualized anatomical models obtained from tissue segmentation, photon transport simulations were conducted to examine intracranial light distribution differences between pediatric and adult groups under standardized stimulation conditions (**Fig. 1D**). The reference coordinates of the light source were predefined in the registration template and subsequently transformed into subject-specific coordinates using the affine matrices generated during T1-weighted MRI segmentation., allowing individualized alignment of the light source position. The light source was modeled as a top-hat beam with a flat intensity profile and uniform forward emission. To accurately capture spatially varying optical properties, voxel-wise updates of optical parameters were applied during the photon transport simulation. This approach enabled precise calculation of photon weight, path length, and scattering angle within heterogeneous media. tissue-specific optical properties at 1064 nm were obtained from previously published literature ^25^ (**Table S1**).

Absorption was modeled using the absorption coefficient *μ^a^* (1/mm), and scattering was modeled using the scattering coefficient *μ^s^*. Directional scattering was described by the anisotropy factor *g*, where *g* = 0 indicates isotropic scattering and *g* = 1 indicates fully forward-directed scattering. Brain tissues typically exhibit *g* ≈ 0.9, which led to the use of the reduced scattering coefficient *μ^s^′* = *μ^s^*⋅ (1−g) in simulations. A uniform refractive index of *n* = 1.37 was assumed for all brain tissues, and cavities or air-filled channels in the tissue atlas were modeled as air. Each simulation launched 1.6 × 10^10^ photons. Photon transport was accelerated using a GPU to parallelize the tracking of photon-tissue interactions, including scattering, reflection and absorption events.

As photons propagated through tissue, their energy attenuated according to the Beer–Lambert law and was deposited locally. The total input energy in each simulation was normalized to 1 J, and energy deposition was recorded for each voxel at 1 × 1 × 1 mm resolution. We selected 10⁻⁶ J/mm³ as the absorption dose threshold (T) under the condition of total input energy normalized to 1 J, as previous work has suggested this level is sufficient to trigger biological effects ^15^. Voxels exceeding this threshold were labeled as activated. This indicates that in both adults and children, a certain cortical absorption volume or proportion must reach the threshold to elicit the desired biological response.

To further quantify intracranial energy distribution, a diffusion–depth curve was computed along the incident beam axis from each subject’s scalp incidence point. Individual curves were obtained first, then averaged within groups to generate group-level profiles (**Fig. 1F**). Considering anatomical differences between children and adults, the proportion of activated GM voxels relative to the total GM volume served as the calibration target. This approach implied that the pediatric group should achieve a GM activation proportion comparable to that of the adult group in order to elicit the desired biological response. The adult group average of this metric, denoted as *R*, served as the reference value. For each pediatric subject *i*, the set of GM voxels was denoted as *G_i_*, with the total voxel count represented as *N_i_*. Under unit-normalized input energy (1 J), the absorbed energy in voxel *j* was represented as *E_j_*. A subject-specific correction factor *k_i_* was determined by iteratively scaling the simulated energy distribution until the child’s GM activation proportion approximated the adult reference. This can be expressed as:

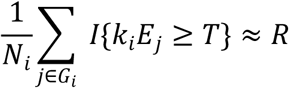

where T is the activation threshold, and I{·} denotes the indicator function.

The group-level calibration coefficient *k* was then obtained by averaging across all valid pediatric subjects. This coefficient was subsequently applied to scale the pediatric stimulation dose in downstream simulations and experimental design.

In Experiment 2, we employed a randomized, triple-blinded, sham-controlled design. Participants were randomly assigned to receive both active and sham sessions, with the order of stimulation counterbalanced and randomized, and a one-week washout period between sessions. As shown in **Fig. 2A**, each session included two post-intervention assessments: the first evaluated immediate effects on selective attention using EEG and cognitive tasks on the same day, while the second assessed short-term clinical outcomes one week later. To ensure blinding, each session was assigned a unique code that was entered into the tLS device by the experimenter. These codes corresponded to either active or sham sessions, but neither the experimenter nor the participants were aware of the specific condition associated with each code. The sequence of codes was randomized by an independent third party who was not involved in any aspect of the assessment process. Blinding procedures were applied to participants, caregivers, clinical evaluators, and data analysts (**Fig. 2C**).

**Fig. 2.**
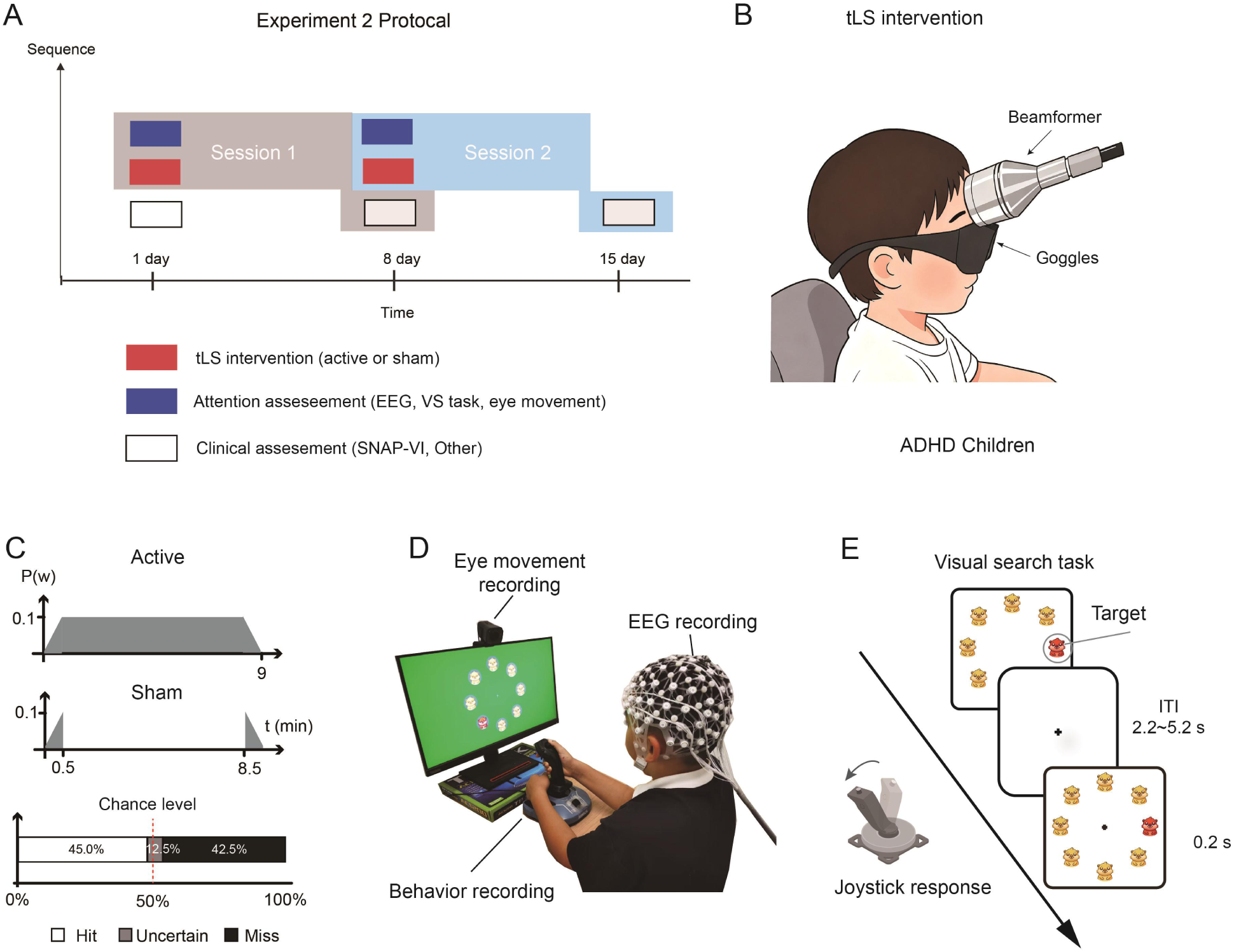
Schematic diagram of the overall experimental design. **A.** Protocol for Experiment 2: Eligible participants determined according to inclusion and exclusion criteria received two tLS sessions (active and sham) in a randomized, triple-blinded design, with a one-week washout period between sessions (session 1 and session 2). **B.** Illustration of the tLS intervention for children with ADHD. **C.** Transcranial light stimulation (tLS) protocol: Active tLS was delivered via a 1064 nm laser targeting the dLPFC for a total duration of 9 min. **D.** EEG setup and attention assessment task illustration. **E.** Response sequence, timeline, and stimulus presented in the visual search paradigm.

As illustrated in **Fig. 1D**, following each session, participants completed the visual search (VS) task (**Fig. 2E**), while their EEG and eye-movement data were recorded synchronously. Participants also underwent standardized attention assessments. For clinical evaluation, both participants and caregivers familiar with them completed the Swanson, Nolan, and Pelham-IV questionnaire (SNAP-IV), The Behavior Rating Inventory of Executive Function (BRIEF), the Sleep Disturbance Scale for Children (SDSC), and the Behavioral Inhibition/Activation System (BIS/BAS) scales. After each intervention, participants completed a validated subjective sensation questionnaire ^26^ designed to capture any potential discomfort. To assess the effectiveness of blinding, participants were asked, after completing both intervention sessions, to indicate which session they believed involved active stimulation and which involved sham stimulation.

This study was approved by the Ethics Committee of Qilu Hospital, Shandong University, and registered with ClinicalTrials.gov (NCT06685601). All procedures followed the ethical standards of the Declaration of Helsinki. Written informed consent was obtained from all participants and their guardians.

In Experiment 3, TD children who met the same inclusion and exclusion criteria as in Experiment 2 were enrolled and completed the VS task (**Fig. 1B**). EEG markers elicited during the VS task in TD children were first analyzed to establish a normative baseline. These reference data were then compared with corresponding EEG markers from children with ADHD following both active and sham tLS sessions. This cross-group analysis was designed to systematically evaluate the neurophysiological effects of tLS modulation.

### Participants

In Experiment 1, a total of 40 participants were enrolled, including 20 adults (mean age = 23.5 years; SD = 2.32, range = 21.4-30.3) and 20 children (mean age = 9.3 years; SD = 1.31, range = 8-14). All participants underwent comprehensive medical screening prior to enrollment. Medical exclusion criteria included a history of traumatic brain injury, neurological disorders, or psychiatric disorders to minimize confounding effects on brain structure and function. MRI–related exclusion criteria included the presence of metallic implants, incompatible medical devices, pregnancy, severe claustrophobia, and tattoos containing metallic pigments could interfere with imaging. Participants meeting any of these criteria were excluded. High-resolution (0.8 mm), 3T MRI scans were obtained for all eligible participants.

In Experiment 2, children with ADHD were recruited from the Pediatric Outpatient Department of Qilu Hospital, Shandong University. As shown in **Fig. 3**, they were evaluated by pediatric psychiatrists to determine whether they met DSM-5 diagnostic criteria for ADHD. The Mini International Neuropsychiatric Interview for Children and Adolescents (MINI-KID, <18 years) was administered to confirm the diagnosis and rule out comorbidities. Eligible participants were also required to demonstrate the ability to cooperate with tLS treatment. Children with an IQ score below 85 on Raven Progressive Matrices, or those undergoing pharmacological or other interventions were excluded. Following outpatient evaluation and screening, 42 children aged 6 to 12 years with ADHD were enrolled, including 33 males and 9 females, (male-to-female ratio = 3.7:1), consistent with the natural incidence rate^27^. The mean age was 8.6 ± 1.4 years. After randomization, two participants withdrew during the intervention and data collection phases. Ultimately, 40 children with ADHD completed all assessments and data collection procedures.

**Fig. 3.**
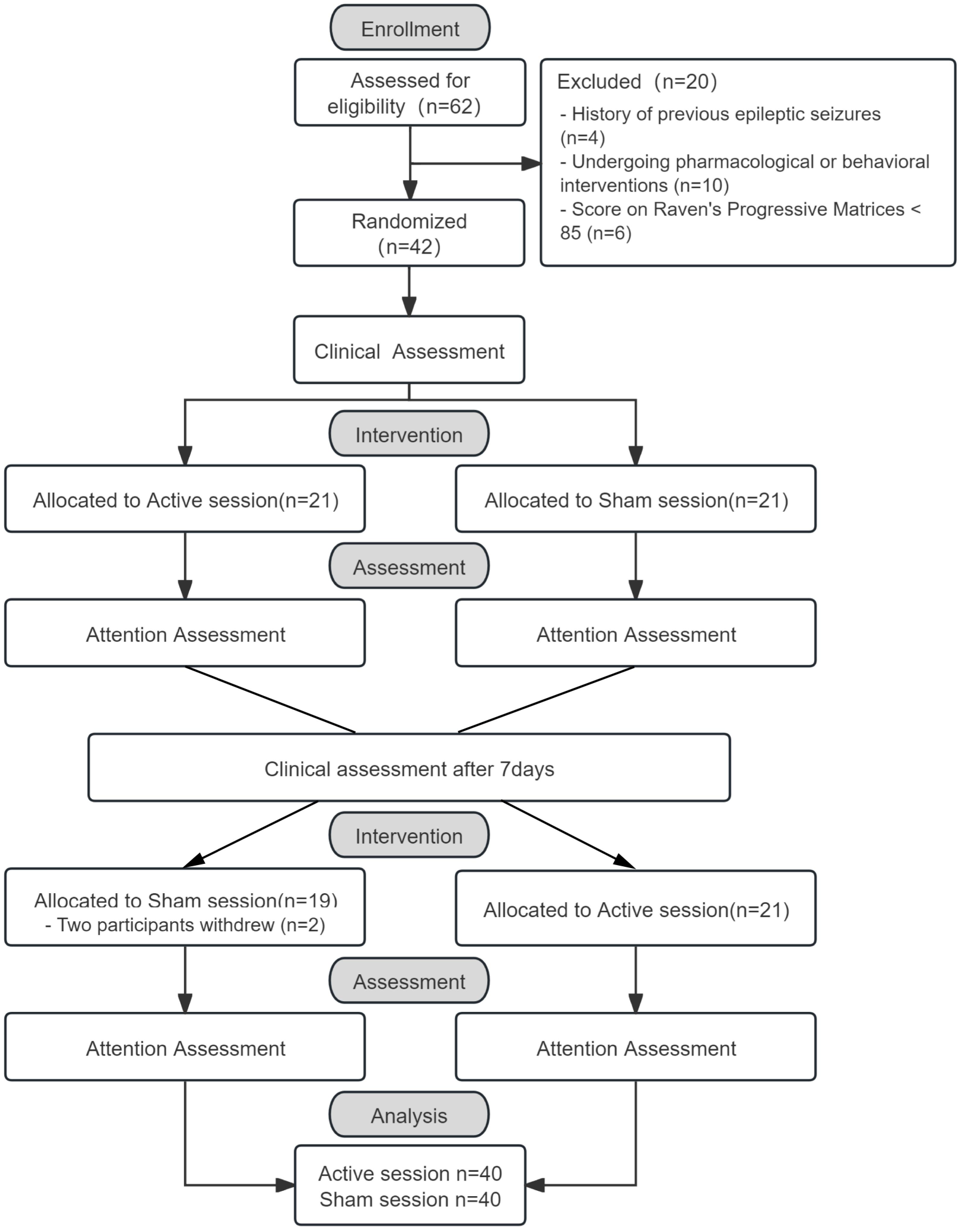
The flow diagram of the study. Participant selection process for children with ADHD in Experiment 2.

In Experiment 3, exclusion criteria included an IQ score < 85 on the Raven’s Progressive Matrices, color vision abnormalities, a personal or family history of neurological or psychiatric disorders, and other relevant abnormalities. The Mini International Neuropsychiatric Interview for Children and Adolescents (MINI-KID) (<18 years) was administered to confirm eligibility. All eligible participants underwent data collection, including EEG and eye-tracking recordings as well as completion of the VS task. Ultimately, 35 TD children completed all data acquisition and met the stringent matching criteria. Matching was performed against the 35 qualified participants from Experiment 2, who were selected using the SNAP-IV as the primary criterion, with strict alignment on age, sex, and IQ scores (**Table 1**).

**Table 1.** Demographic characteristics and diagnostic information of study partici pants.

| Characteristics | ADHD<sup>a</sup> | TD | P value |
| --- | --- | --- | --- |
| Age (years, Mean ± SD) | 8.64 ± 1.46 | 8.76 ± 1.25 | 0.723<sup>b</sup> |
| Sex (Female/Male) | 7/28 | 9/26 | 0.569<sup>c</sup> |
| **ADHD-Subtype** | **N (%)** | / | / |
| Combined (n) | 9 (25.71 %) | / | / |
| Hyperactive/impulsive (n) | 2 (5.71 %) | / | / |
| Inattentive (n) | 24 (68.57 %) | / | / |
| Raven's Progressive Matrices | 114.83 ± 10.78 | 118.83 ± 14.79 | 0.098<sup>d</sup> |

Note. <sup>a</sup> The number of eligible participants remaining in SNAP-IV after data verification and screening. <sup>b</sup> p Values calculated by 2-sample t test. <sup>c</sup> p Values calculated by $\chi^2$ test. <sup>d</sup> P values were calculated using the Mann-Whitney U nonparametric test due to non-normal distribution of data in the TD group.

### tLS Protocol

A diode-pumped solid-state laser with a linewidth of approximately ± 0.1 nm was used in Experiment 2. Based on the adult reference energy of 1500-1600 J and the group-level calibration coefficient (0.4) derived from Experiment 1, the recommended stimulation dose for the pediatric group was set to approximately 600-640 J. The measured uniform laser beam area was 12.57 cm^²^ (diameter = 4 cm), and the irradiance (power density) was set to 100 mW/cm^²^ (250*0.4 = 100 mW/cm²). The total exposure time was 8 min, resulting in a total energy of 640 J. At these parameters, the emitted energy remained well below the maximum permissible exposure for the skin, ensuring no detectable physical damage or thermal effects. The probe used in this study was handheld and positioned on the participants’ right frontal lobe, centered at FP2, with EEG electrodes placed according to the international 10-20 system. During the active tLS session, participants received a 8-min intervention at an intensity of 100 mW/cm^²^, with the power gradually ramped up over 30 s. During the sham session, stimulation was delivered only during the first and last 30 s, using the same wavelength and irradiance, with no stimulation during the intervening 8 min, the laser power was reduced to zero during this period. The two 0.5-min stimulation created a subjective experience similar to the active stimulation, minimizing potential biases in participant responses.

### Attention Assessment Task

The study employed a VS task ^28^. Experimental stimuli were presented on a 27-inch monitor with a resolution of 1920 × 1080 pixels. Participants were seated comfortably at a distance of 58 cm from the screen, with seat height adjusted individually to ensure that the visual axis was aligned with the center of the display. The task procedure is illustrated in **Fig. 2E**. At the beginning of each trial, participants fixated on a central cross. After 2 s, a search matrix containing eight circular groundhogs (1.7° × 1.7°, brightness 13.5 cd/m^²^) was displayed for 0.2 s. Participants were instructed to determine the direction of the target (red groundhog) relative to the fixation point. Once the target appeared, they used a joystick to rapidly move in the direction of the target and confirmed their response by pressing the joystick confirmation button (**Fig. 2E**). The task consisted of six blocks, each containing 46 trials, for a total of 276 trials. Participants were allowed a maximum rest period of 6 min between blocks.

### Eye tracking

Eye gaze was calibrated and tracked using online software, that applies deep learning–based modeling for webcam-based eye-tracking (https://github.com/Labvanced). Gaze recordings were collected while participants were seated approximately 58 cm from the screen center. At the beginning of each block, a 15-point, single-pose calibration was administered, with an error threshold set at 15% of the screen size. Calibration typically required about 45 s. Each calibration point was paired with an animal icon and sound to maintain children’s attention. If the error threshold was exceeded, calibration was repeated until the criteria were met, and experimental blocks only began once calibration was successful. During task trials, head pose alignment was continuously monitored, gaze data were sampled at 5 Hz, and webcam-based head and face tracking remained active.

### EEG recording and processing

EEG data were recorded using a 64-channel saline-based electrode cap following the international 10-20 system, in combination with a SynAmps2 amplifier and the Curry 9.0 package (Compumedics Neuroscan, USA). During data acquisition, electrode impedances were maintained below 50 kΩ. Data were sampled at 1000 Hz, and a bandpass filter was applied with a passband ranging from 0.01 to 400 Hz.

Preprocessing was performed in MATLAB using the EEGLAB toolbox ^29^. Data were downsampled to 250 Hz and bandpass-filtered between 0.5 and 30 Hz. Prior to independent component analysis (ICA), segments contaminated by excessive artifacts were manually removed, and electrodes with poor signal quality were identified and interpolated. Independent components associated with vertical eye movements were manually identified and removed, after which an automatic artifact rejection algorithm was applied to exclude EEG epochs in which voltage exceeded ±100 μV at any electrode.

ERP preprocessing was performed in MATLAB using the ERPLAB toolbox ^30^. Data were re-referenced to the average reference and epochs from -200 ms to 400 ms relative to stimulus onset were extracted. The N2pc components was derived from difference waves by subtracting ipsilateral from contralateral waveforms at target-related electrodes. Because the overall stimulus energy was bilateral, this subtraction removed most unrelated ERP components, leaving a clear N2pc signal. N2pc amplitudes were measured at the PO7/PO8 electrode sites, as reported in previous studies ^31^. Amplitudes were calculated as the mean voltage within three samples centered on the peak, determined in the 220-320 ms post-stimulus window. Group comparisons of N2pc amplitudes between children with ADHD (under sham and active sessions) and the TD control group were conducted using independent-samples t-tests.

### Inverted Encoding Model (IEM) Analysis

To reconstruct location-specific channel tuning functions (CTFs) from EEG topographies, we employed an inverted encoding model (IEM) approach ^32^. The IEM was used to track the time course of spatially specific alpha-band (8-12 Hz) activity across electrodes. The model assumes that the observed power at each electrode (sampled at each time point) reflects the weighted sum of responses from eight spatially selective channels. Each channel’s response was modeled using a half-sinusoid raised to the ninth power:

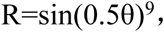

Where θ represents the angular location (0°, 45°, 90°, 135°, 180°, 225°, 270°, 315°, 360°), and R is the corresponding channel response.

EEG signals were segmented into 1000 ms epochs around target onset (-200 to 800 ms). The data were bandpass filtered between 8 and 12 Hz using the eegfilt function from EEGLAB^33^, and instantaneous power estimates were obtained via Hilbert transformation (MATLAB, MathWorks, Natick, MA). The IEM was then applied to each time point in the alpha-band power data.

Artifact-free epochs were divided into training (B_1_) and testing (B_2_) datasets for each participant. B_1_ and B_2_ represent matrices of power at each electrode for trials in the training and testing sets, respectively. Channel responses (C_1_) for the training set were determined based on the hypothetical tuning functions for each spatial location. The relationship between electrode-level power (B^1^) and channel responses (C_1_) was modeled using a general linear model:

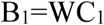

Where W is the weight matrix mapping channel space to electrode space. The weight matrix was estimated using least-squares regression:

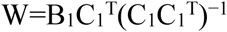

In the testing phase, channel responses (C_2_) were estimated from the observed electrode-level test data (B2) using the estimated weight matrix:

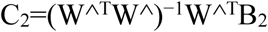

The estimated responses (C_2_) were realigned so that the center channel corresponded to the channel tuned to the actual stimulus location (θ). Aligned responses were averaged across trials to generate the final CTFs. The slope of the CTF at each time point was computed and used as an index of spatial attention deployment toward the distractor.

### Clinical Assessments

As shown in **Fig. 2A**, multiple questionnaires were collected at one-week intervals following each tLS intervention. Both participants and their caregivers completed assessments. The SNAP-IV, SDSC, and BRIEF questionnaires were completed by the participants’ primary caregivers, who were most familiar with the children’s behavior. Prior to data analysis, rigorous verification procedures were applied. Questionnaires with poor response quality, missing data, or logical inconsistencies were either imputed or excluded. The number of valid responses for each questionnaire was as follows: SNAP-IV (n = 35), SDSC (n = 33), and BRIEF (n = 27).

A total of three assessments were conducted on Days 1, 8 and 15. For the first assessment, caregivers were instructed to evaluate participants’ typical behavior, whereas for the second and third assessments, they were asked to reflect on behavior during the week following the intervention. The Day 1 assessment served as the baseline and the results of Day 8 and Day 15 were used to evaluate post-intervention performance for the active and sham sessions, respectively. The BIS/BAS scales in contrast to the caregiver-reported questionnaires, were completed by the participants themselves. The Day 8 and Day 15 assessments reflected behavior following the active and sham sessions, while Day 1 represented the baseline level. For the inter-group comparisons between active and sham sessions, paired t-tests were used for normally distributed data, and Wilcoxon signed-rank tests were applied when data deviated from normality. A significance threshold of p < 0.05 was adopted for all statistical evaluations. Because multiple sub-scores were analyzed, False Discovery Rate (FDR) correction was applied to control the rate of Type I errors.

## Results

### Simulation of Dosimetry

As shown in **Fig. 1A**, Experiment 1 enrolled 20 ADHD children, each undergoing high-resolution MRI acquisition, tissue segmentation, and Monte Carlo photon transport simulation. Representative examples from each group are displayed in **Fig. 1B**, showing raw T1-weighted images alongside CAT12-based segmentation into major tissue classes, including GM, WM, CSF, skull, and scalp. Group-level comparisons (**Fig. 1C**) revealed significant anatomical differences between children and adults, particularly in GM, WM, and skull proportions. These structural differences are expected to influence intracranial photon propagation.

Monte Carlo simulations confirmed the influence of anatomical structures on photon distribution. Energy deposition maps (**Fig. 1D**) demonstrated intracranial differences between the two groups under identical incident dose conditions. As shown in **Fig. 1E**, the number of voxels exceeding the activation threshold was higher in children, particularly within GM, indicating disproportionate energy delivery relative to adults. To further characterize photon diffusion along the depth axis, the group-averaged diffusion area–depth relationship was computed (**Fig. 1F**). The pediatric group exhibited larger activated areas across multiple depth planes, with more pronounced differences in GM activation. These findings are attributed to thinner skull and distinct tissue composition in children (**Fig. 1B**).

A group-level calibration coefficient (k = 0.40) was derived from individual results. After applying this correction, the GM activation proportion in the pediatric group was reduced and more closely aligned with the adult reference value, confirming the feasibility of parameter transfer across populations

### Blinding Assessment and Subjective Sensory Evaluation

The blinding assessment confirmed that participants remained unaware of their treatment allocation, with no evidence of expectancy bias. Among the 40 participants, guessing accuracy (17 correct [42.5%], 18 incorrect [45%], and 5 uncertain [12.5%]) did not differ significantly from chance (**Fig. 2C**).

In the subjective sensation questionnaires completed after each intervention, only a few participants reported mild warmth or fatigue. All ratings were mild or below and any sensations of discomfort subsided within minutes after stimulation ceased.

### Changes in Clinical Assessments

As shown in **Table 2**, SNAP-IV results demonstrated significant improvements following the active session compared with the sham session in both the total score (t = -2.659, p = 0.012, p^FDR^ = 0.036, Cohen’s d = -0.450, two-tailed) and the inattention subscale (t = -2.495, p = 0.018, p^FDR^ = 0.036, Cohen’s d = -0.422, two-tailed). No significant differences were observed between sessions in hyperactivity-impulsivity (t = -1.506, p = 0.141, p^FDR^ = 0.188, Cohen’s d = -0.254, two-tailed) or oppositional defiance (t = 1.039, p = 0.306, p^FDR^ = 0.306, Cohen’s d = -0.176, two-tailed) (**Table 2**).

**Table 2.**
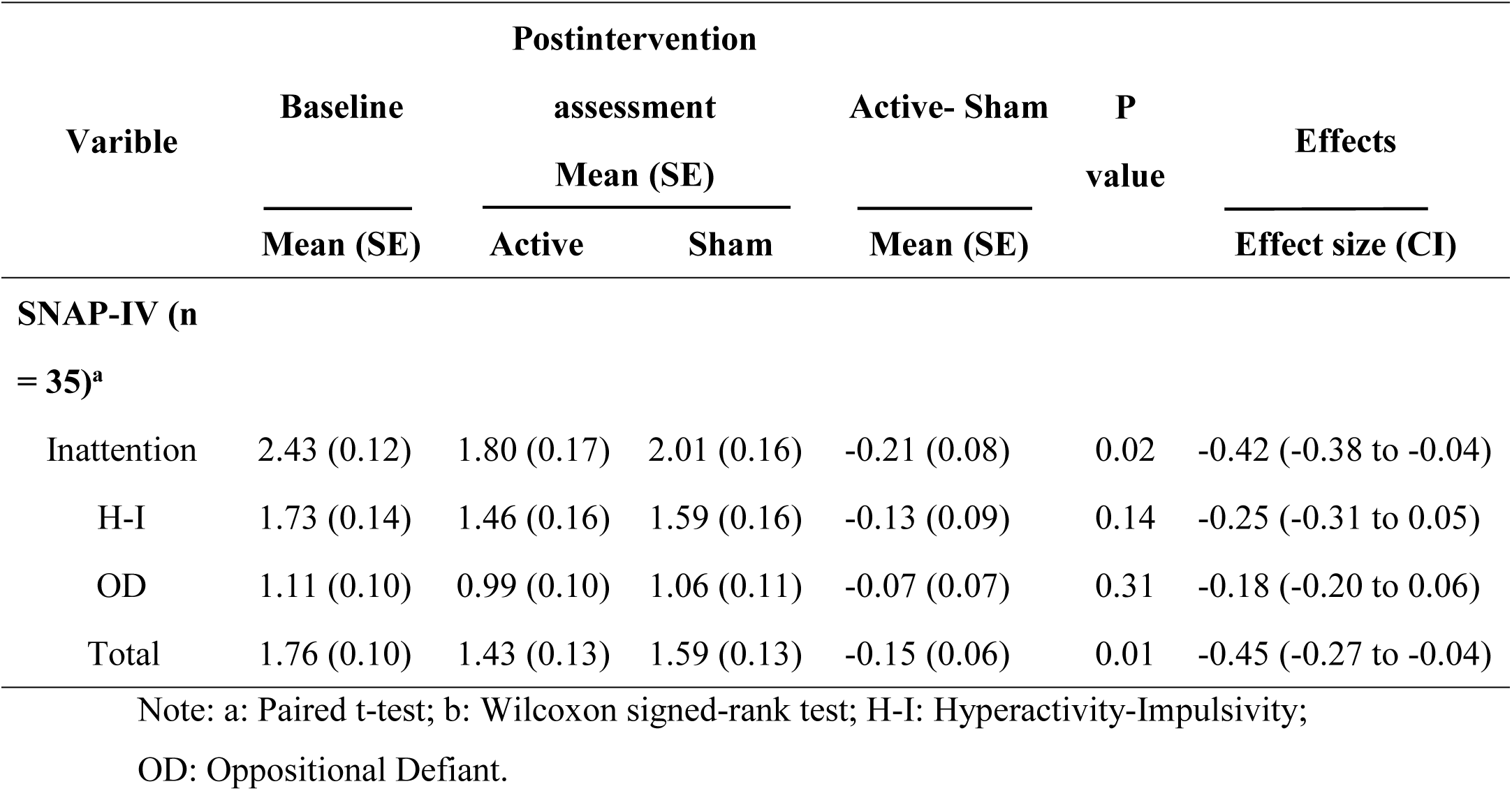
Statistical Results for the SNAP-Ⅳ Rating Scale Postintervention.

In the SDSC (**Fig. S2B**), significant improvement was observed in the Behavioral Sleep Problems (BSP) subscale following active stimulation (Z = -2.143, p = 0.032, p^FDR^= 0.224, r = -0.373). No significant differences were found between sessions in the total score or in the Sleep Breathing Disorder, Arousal Disorder, Sleep-Wake Transition Disorder, Excessive Daytime Somnolence, and Sleep Hyperhidrosis subscales (all p-value > 0.147) (**Table S2**).

The results of the BRIEF scale are presented in **Fig. S1B**, The active stimulation group showed significant improvements compared with the sham group in the Global Executive Composite (t = -2.104, p = 0.045, p^FDR^= 0.099, Cohen’s d = -0.405, two-tailed), Metacognition Index (t = -2.726, p = 0.011, p^FDR^= 0.061, Cohen’s d =-0.525, two-tailed), Working Memory (t = -2.886, p = 0.008, p^FDR^= 0.061, Cohen’s d = 0.555, two-tailed), Self-Monitoring (t = -2.199, p = 0.037, p^FDR^= 0.099, Cohen’s d = 0.423, two-tailed), and Materials Organization (Z = -2.164, p = 0.030, p^FDR^= 0.099, r = -0.416). In the domains of Planning and Organization (t = -1.831, p = 0.079, p^FDR^= 0.124, Cohen’s d = 0.352, two-tailed) and Initiation (Z = -1.889, p = 0.059, p^FDR^= 0.108, r = -0.364), there was a trend toward improvement following active stimulation, approaching marginal significance. However, no significant differences were observed between groups in Inhibition, Shifting, Emotional Control, or the Behavior Regulation Index (all p-value > 0.209) (**Table S2**).

The BIS/BAS results completed by the participants are presented in **Fig. S1A**. The BAS total score following active stimulation was significantly higher than that observed after sham stimulation (t = 2.083, p = 0.046, p^FDR^ = 0.115, Cohen’s d = 0.374, two-tailed). In contrast, the BIS total score showed a trend toward marginal significance between the two conditions (t = -1.850, p = 0.074, p^FDR^ = 0.123, Cohen’s d = -0.332, two-tailed). Among the three BAS subscales, a significant increase was observed in Reward Responsiveness after active stimulation (Z = -2.353, p = 0.019, p^FDR^ = 0.095, r = -0.422), whereas no significant between-group differences were found for the Fun-Seeking and Drive subscales (all p-value > 0.319) (**Table S2**).

### Selective Attention Behavioral Results

Consistent with previous studies ^34^, reaction times in the VS task were calculated as a behavioral index of selective attention. Performance was compared between sham and active sessions within the ADHD group, as well as between ADHD and TD groups. Within the ADHD group, reaction times were significantly faster in the active session than in the sham session (t = -2.161, p = 0.038). In the between-group comparison, ADHD participants showed significantly slower reaction times than TD group under the sham session (t = 1.489, p = 0.014), whereas no significant difference was observed under the active condition (t = 2.521, p = 0.141). These results suggest that, without intervention, children with ADHD exhibited a processing speed deficit that was alleviated following active tLS.

### Changes in ERP of Selective Attention

We next examined ERPs during the VS display and calculated the N2pc component as an index of selective attention. The N2pc appeared as a negative deflection in the ERP waveform at the visual cortex contralateral to the target. The average N2pc amplitude was measured in the 245-285 ms window at PO7/8 (**Fig. 4B**). As shown in **Fig. 4D**, Within the ADHD group, N2pc amplitude differed significantly between active and sham sessions (t = -2.14, p = 0.042). In the between-group comparison, ADHD participants showed a marginally significant reduction in N2pc amplitude compared with TD controls in the sham session (t = -2.00, p = 0.052), whereas no significant difference was observed in the active session (t = -0.22, p = 0.831). These findings indicate that ADHD participants displayed a selective attention deficit in the absence of intervention, which was no longer evident following tLS. For N2pc latency (**Fig. 4E**), no significant difference was observed between active and sham sessions within the ADHD group (t = 0.867, p = 0.394), nor between ADHD and TD groups under either active (z = -0.91, p = 0.365) or sham stimulation (t = 0.38, p = 0.706). In the correlation analysis (**Fig. 4M**), a smaller negative difference in N2pc amplitude between active and sham stimulation was positively associated with higher scores on the inattention subscale of the SNAP-IV, indicating that more positive N2pc differences were linked to more pronounced inattentive symptoms.

**Fig. 4.**
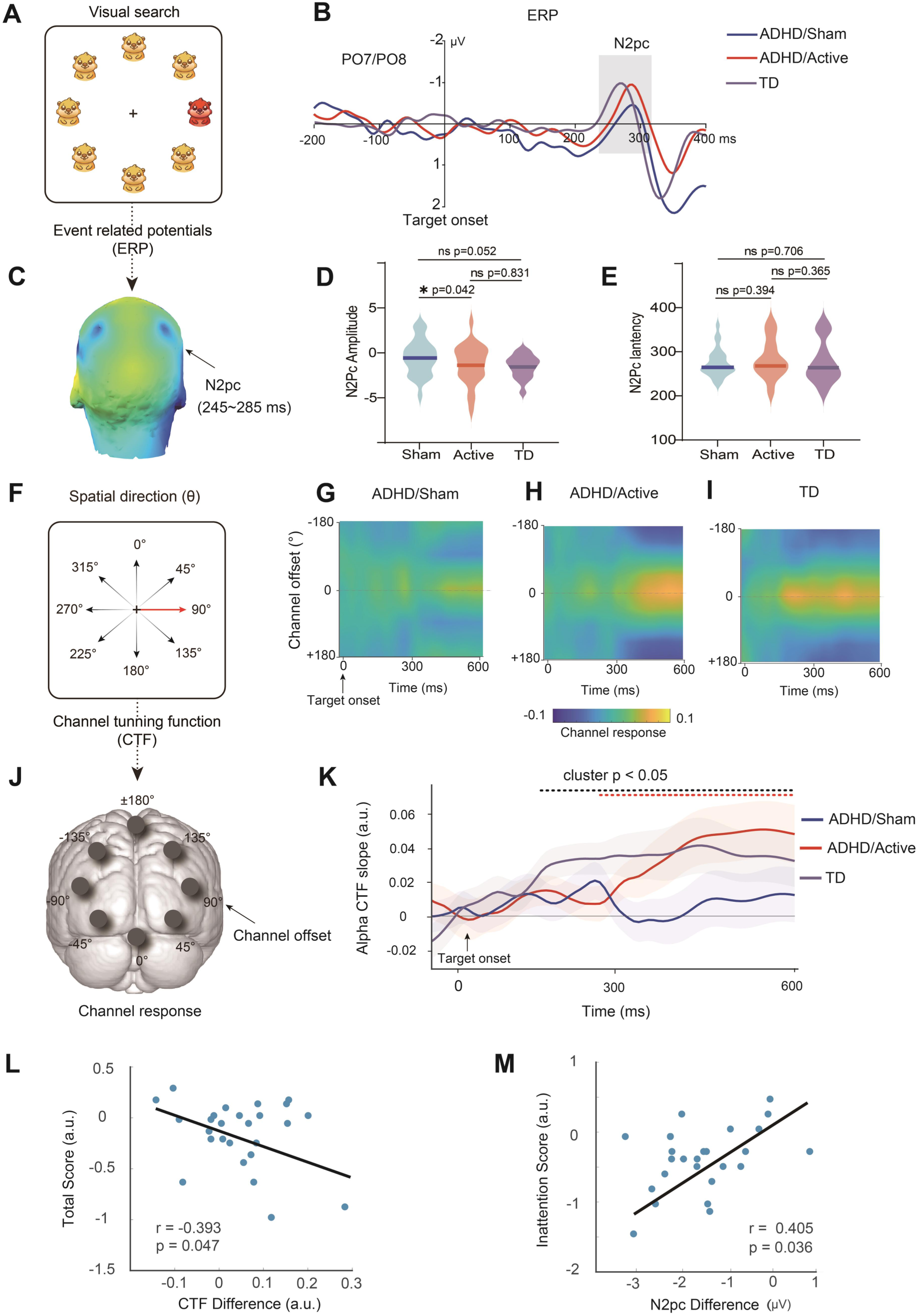
Effects of tLS on selective attention in children with ADHD. **A.** A visual search stimulus while ERPs were recorded to capture neural responses to target (red). **B.** ERP waveforms at PO7/PO8 electrodes. The N2pc component emerged approximately 245–285 ms after target onset. Clear N2pc responses were observed in TD (purple) and ADHD/Active (red) groups, while the ADHD/Sham (blue) group showed a reduced response. **C.** Scalp topography of voltage distribution during the N2pc window (245–285 ms), highlighting lateralized posterior activation linked to attentional allocation. **D.** Between-group comparisons of N2pc amplitude across the TD (purple), ADHD/Active (red), and ADHD/Sham (blue) groups during the N2pc window (245–285 ms). **E.** Between-group comparisons of N2pc latency across the TD (purple), ADHD/Active (red), and ADHD/Sham (blue) groups during the N2pc window (245–285 ms). **F.** Schematic of channel tuning function (CTF), illustrating neural channels respond to different spatial directions (θ) during attentional processing. **G–I.** Time-resolved CTF maps showing channel responses by channel offset across time in ADHD/Sham (G), ADHD/Active (H), and TD (I) groups. These maps illustrate group differences in spatially selective alpha-band responses. **J.** Topographic layout of channel offsets used to compute the CTF, depicting spatial tuning across EEG channels. **K.** Time course of alpha-band CTF slope, indicating attentional selectivity over time. TD (black) and ADHD/Active (red) groups showed increased tuning following target onset, whereas the ADHD/Sham (blue) group showed no significant increase. A significant group effect was observed in the 320–600 ms cluster (p < 0.05). **L.** Association between changes in SNAP-IV total scores and differences in CTF values after active versus sham stimulation. **M.** Association between changes in SNAP-IV inattentive subscale scores and differences in N2pc amplitudes after active versus sham sessions.

### Alpha Channel Tuning Function of Target

To capture spatial attention with high spatiotemporal resolution, we employed an IEM to reconstruct the location of selectively attended targets from alpha-band power patterns recorded with scalp EEG. As shown in **Fig. 4F-J**, this method yields channel tuning functions (CTFs), which describe the distribution of alpha power across eight idealized spatial channels. The central channel was aligned with the target direction (e.g., 90° red target in **Fig. 4F**), and channel offsets was defined as the angular difference between the center and other channels (e.g., 90° offset in **Fig. 4J**, right panel). For cross-trial comparison, all estimated CTFs were circularly shifted so that the target location was centered at 0° on the channel offset axis, resulting in a standardized representation with offsets ranging from -180° to 180°. Figure 4G–I show time-resolved CTFs (0 to 600 ms relative to target onset) for ADHD participants under sham session, ADHD participants under active session, and TD group. Spatial selectivity was quantified by computing the CTF slope over time (**Fig. 4K)**, where steeper slopes reflect greater tuning precision. The results demonstrate that alpha-band CTFs captured the dynamic profile of spatial attention. The TD group exhibited a significant increase in CTF slope following target onset (164-600 ms; permutation test, cluster-corrected p < 0.05, two-tailed). The ADHD active session showed a similar trend from 320 to 600 ms (cluster p < 0.05, two-tailed), suggesting enhanced attentional selectivity with tLS. In contrast, the ADHD sham session did not show a comparable increase during this period. A significant group effect was also observed between 435 and 600 ms post-target (p < 0.05), indicating early differences in attentional allocation across groups. In the correlation analysis between CTF differences and SNAP-IV total scores (**Fig. 4L**), larger CTF differences were associated with lower SNAP-IV total scores, indicating that greater CTF differences corresponded to less pronounced inattentive symptoms.

## Discussion

This study aimed to enhance spatial selective attention in children with ADHD by applying tLS at 1064 nm over the right DLPFC. Clinical observations during the following week revealed a significant reduction in clinical rating, particularly in the inattention domain during the active session. Furthermore, the immediate effects on attention performance were assessed following tLS. Behavioral performance on the selective attention task, together with neural biomarkers (N2pc amplitude and CTF responses), showed significant improvement after active tLS compared with the sham session. Changes in EEG markers reflecting attentional enhancement after tLS were significantly correlated with subsequent reductions in inattention symptoms, suggesting a neurophysiological basis for clinical improvement in ADHD.

Non-invasive brain stimulation techniques have been applied in the rehabilitation of neurodevelopmental disorders in children and adolescents, including rTMS, tDCS and tLS. In pediatric studies, strategies for determining stimulation dosage vary considerably across these modalities. TMS typically defines stimulation intensity as a percentage of the individual’s resting motor threshold ^35,36^, which can be measured rapidly during the experiment and adjusted in real time for individualized dosing. In contrast, tDCS often adopts a uniform low-intensity protocol (e.g., 1–2 mA), with pediatric applications generally applying slight reductions from adult parameters ^37^. Unlike these approaches, the depth and distribution of photon propagation in tLS are influenced not only by light source parameters but also by anatomical factors such as skull thickness, tissue composition, and tissue-specific optical properties. To date, only a limited number of studies have investigated the application of tLS in pediatric populations, most of which have used LED-based devices ^37^. These devices typically feature relatively low irradiance and limited energy delivery capacity, with notable variation in form factors and emission characteristics. Consequently, pediatric dosing in these studies has generally been determined conservatively or selected empirically to provide a fixed experimental dose, rather than based on quantitative modeling. Given the substantial inter-individual variability in light propagation and tissue absorption, this highlights the importance of incorporating subject-specific anatomical modeling and photon transport simulations to establish optimized dosing strategies. Such approaches are essential to ensure both safety and therapeutic efficacy in individualized tLS applications, particularly for pediatric populations.

In Experiment 1, we found that anatomical differences between children and adults—particularly in skull thickness and tissue composition had a clear impact on transcranial light distribution, as demonstrated by the Monte Carlo photon transport simulations. Under identical incident energy conditions, the pediatric group exhibited a significantly greater number of GM voxels exceeding the activation threshold (10⁻⁶ J/mm³) compared with the adult group (**Fig. 1F**). These findings suggest that directly applying adult stimulation parameters to children may result in disproportionate energy deposition in GM increasing the risk of overstimulation or producing a bidirectional dose–response effect. To address this, GM activation ratio was selected as the calibration target, with the goal of aligning the pediatric group’s energy distribution with that of the adult group to elicit a comparable biological response. Results showed that, when total input energy was normalized to 1 J and the group-level coefficient was set to 0.40, the GM activation ratio in the pediatric group decreased and closely approached the adult reference level. This indicates that the proposed calibration method effectively reduced energy distribution bias caused by anatomical differences, providing a quantitative basis for transferring dose parameters to subsequent experiments.

These findings should be considered in the context of theoretical models of attention impairments in ADHD ^38^. Compared with TD children, the ADHD group exhibited deficits in selective attention, as indicated by prolonged EEG latencies and reduced component amplitudes ^39^. This perspective is further supported by cross-experimental comparisons (**Fig. 4**), which showed that, during the sham session in Experiment 2, EEG markers of ADHD participants consistently differed from those of TD children in Experiment 3.

Following tLS treatment, ADHD participants demonstrated significant improvements in both the SNAP-IV total score and its inattention subscore compared with the sham session. The amplitude of the N2pc component was also significantly enhanced after the active session, approaching the level observed in TD children. These results suggest that tLS may strengthen top-down attentional control over spatial information processing, thereby improving the ability to selectively attend to relevant stimuli, as reflected by a sharper CTF slope (**Fig. 4K**). Previous studies have similarly reported that enhancing attentional selectivity promotes better retention of task instructions and more efficient motor responses ^40^.

As shown in **Fig. S1D**, children with ADHD exhibited significantly shorter response times following active tLS compared with sham session (p < 0.038). This acceleration in both information processing and motor execution indicates enhanced cognitive-motor efficiency, leading to improved behavioral performance. Results from the BRIEF also revealed significant improvements in working memory, metacognition, and other executive functions after active stimulation, consistent with previous findings in both healthy adults and patients with ADHD ^16,19^. Notably, tLS also produced significant improvements in the total BAS score and its Reward Responsiveness subscale. This suggests that stimulation of the prefrontal cortex may modulate neural circuits underlying reward and motivation ^41^, helping children experience greater positive affect from feedback and thereby reducing frustration or oppositional behaviors linked to insufficient motivation. Based on neuroimaging studies of atypical brain regions in ADHD ^42^, the mechanism underlying attention improvements with tLS may involve enhanced excitability of the right dlPFC, which strengthens executive control over the selective encoding of spatial information in parieto-occipital regions.

In Experiment 2, we evaluated the effects of tLS by comparing attentional performance before and after intervention and further contextualized these changes relative to baseline data from TD participants in Experiment 3. This comparison allowed us to estimate the extent to which tLS modulated performance toward normative levels. Interestingly, although only short-term offline effects (< 30 min post-stimulation) were assessed in Experiment 2, the observed improvements in EEG measures approached levels comparable to those of the TD group, suggesting that tLS may effectively reduce clinical symptoms in children with ADHD.

Compared with traditional home-based NIBS approaches, which typically require multiple weeks of intervention to produce measurable clinical improvements, tLS may provide a more rapid and tolerable alternative. Its favorable safety profile, particularly compared with tDCS, (which often causes skin irritation), supports its potential suitability for pediatric populations and wearable applications ^43^.

The inclusion of TD participants in Experiment 3 provided an objective reference point, confirming that tLS-induced changes in attentional performance and EEG biomarkers occurred in a direction consistent with potential clinical benefit. Moreover, deviations of EEG markers from TD norms may serve as predictors of treatment responsiveness, supporting the promise of individualized, developmentally informed tLS strategies in future applications.

This study has several limitations that should be acknowledged. First, the single-session intervention design may amplify transient stimulation effects, and the observed outcomes could have been influenced by confounding variables. Second, the relatively small sample size and exploratory nature of the study preclude definitive conclusions regarding the clinical efficacy of tLS for ADHD. Large-scale studies incorporating repeated sessions and long-term follow-ups will be necessary to determine whether the observed effects translate into clinically meaningful improvements. Third, we did not differentiate between ADHD subtypes (e.g., inattentive, hyperactive-impulsive, or combined presentation), which may present distinct neural profiles and treatment responses. Future research should recruit larger and more diverse samples to examine potential subtype-specific effects of tLS.

The present study demonstrates that tLS using near-infrared II wavelength light over the right dlPFC can enhance spatial selective attention in children with ADHD. These findings have important implications for future research, including the optimization of tLS protocols for early intervention in pediatric neurodevelopmental disorders (e.g., autism spectrum disorder). Our results suggest that tLS may serve as an alternative therapeutic approach for modulating selective attention deficits and inattention symptoms in ADHD. Furthermore, this study provides preliminary empirical support for the potential efficacy of repeated tLS sessions in ADHD. Additionally, our findings indicate that this protocol may also be relevant for other neurodevelopmental and psychiatric disorders characterized by attentional impairments, such as ASD, dyslexia, and depression, thereby offering a possible intervention for shared clinical symptoms.

## Data Availability

All data produced in the present study are available upon reasonable request to the authors

## Acknowledgments

This work was supported by OpenResearch Fund of Beijing University of Posts and Telecommunications and the Qilu Special Fund for Pediatric Discipline Development.

## Author contributions

C.Z. and A.C. supervised the study. Y.Z., Y.L., K.Z., and C.Z. processed data and prepared the manuscript. C.Z., Y.L., Z.L., and Y.H. designed the experiment. Y.L., H.T., Y.H., and H.J. collected data. Z.C. and Y.H. provided analytical support and revised the manuscript.

## Data availability

All data produced in the present study are available upon reasonable request to the authors.

