## Supplemental Data 1 for "Transcranial Light Stimulation Improves Selective Attention in Children with ADHD"

### Supplemental information

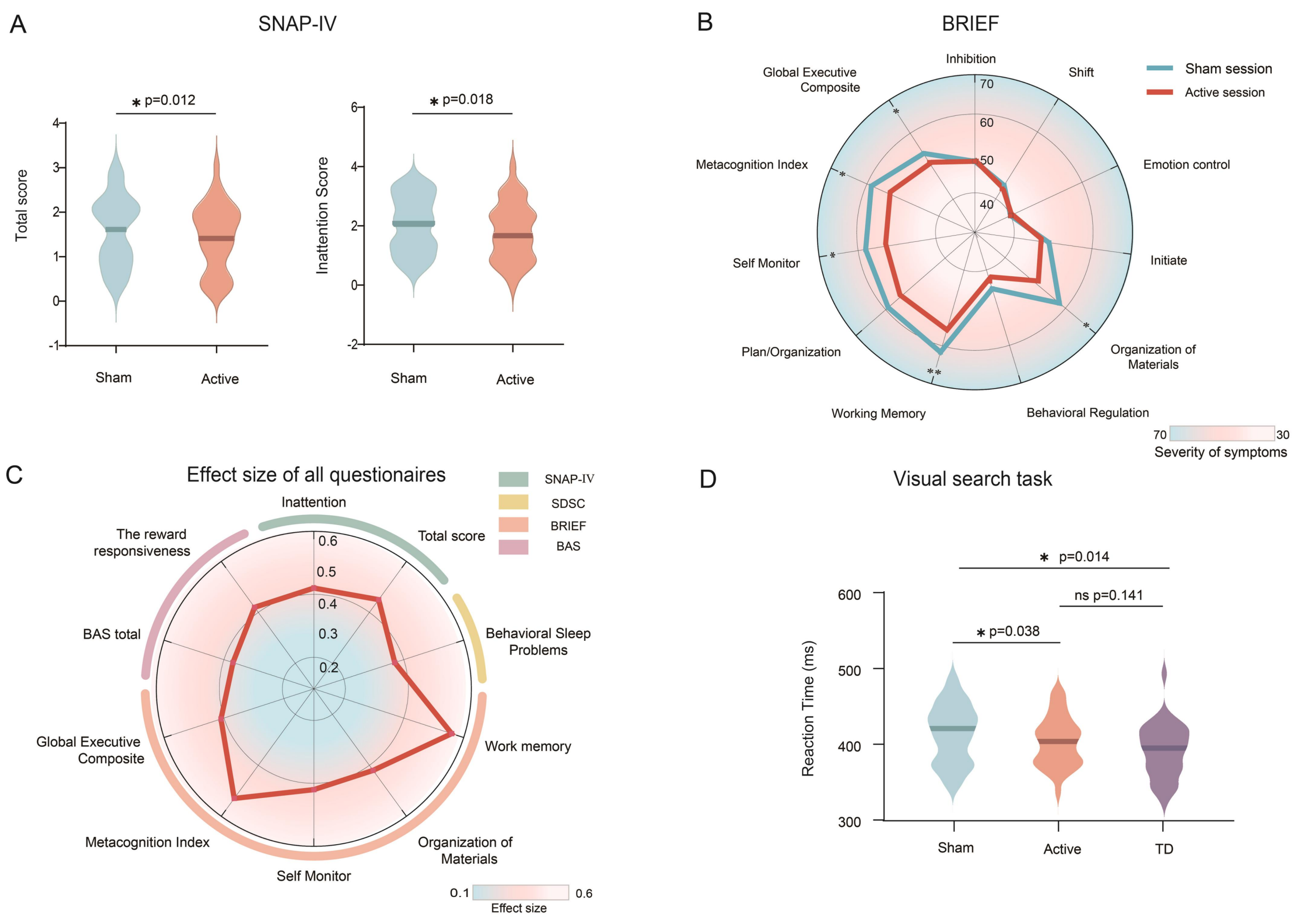


**Fig. S1. Between-group comparisons of major clinical assessment scores and corresponding effect sizes. A.** Total scores and subscale results of the SNAP-IV. **B.** T-scores of BRIEF domains and clinical scores. Subscales including Inhibit, Shift, Emotion Control, Initiate, Organization of Materials, and Behavioral Regulation Index showed normal distributions and were analyzed using paired-samples t-tests. Remaining data exhibited non-normal distributions and were analyzed using non-parametric tests. **C**. Effect sizes (Cohen's d) of tLS treatment across the 10 outcome measures. **D.** Violin plots of reaction times in the visual search task for ADHD/Sham (blue), ADHD/Active (red), and typically developing (TD， purple) groups. Horizontal lines indicate median reaction times. Asterisks denote statistically significant group differences ($p < 0.05$), and "ns" indicates non-significance.

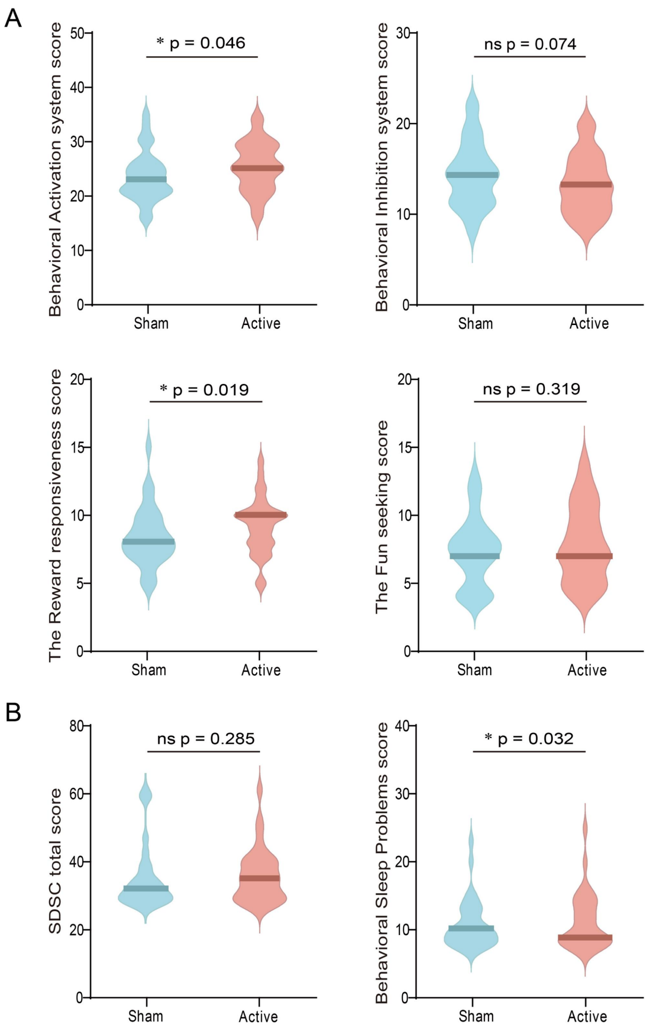


**Fig. S2. Between-group comparisons of clinical assessment scores. A.** Total scores of the BAS/BIS scales and subscale scores of the BAS. **B.** Total score of the SDSC and score of its BSP subscale.

**Table S1. Optical parameters of brain tissues at 1064 nm.**

| | 1064 nm | | | |
|---|---|---|---|---|
| | $\mu_s$ | $\mu_s'$ | $\mu_a$ | $g$ |
| WM | 30 | 3.6 | 0.105 | 0.88 |
| GM | 5.9 | 0.53 | 0.053 | 0.91 |
| CSF | 0.09 | 0.01 | 0.0144 | 0.89 |
| Skull | 14.6 | 1.61 | 0.019 | 0.89 |
| Skin | 18.4 | 2.03 | 0.017 | 0.89 |

Note. $\mu_s$: scattering coefficient (1/mm); g: anisotropy factor; $\mu_s'$: reduced scattering coefficient (1/mm); $\mu_a$: absorption coefficient (1/mm).

**Table S2. Statistical Results for Clinical Rating Scales in ADHD**

| Varible | Baseline Mean (SE) | Postintervention assessment Mean (SE) | | Active- Sham | Effects | P value |
|---|---|---|---|---|---|---|
| | | Active | Sham | Mean (SE)/ Median | Effect size(CI) | |
| **Sleep Disturbance Scale for Children (SDSC) (n= 33)[b]** | | | | | | |
| Behavioral Sleep Problems (BSP) | 11.76 (0.69) | 11.30 (0.71) | 10.70 (0.63) | -1.00 | 0.37 (-2 to 4) | 0.03 |
| Sleep Breathing Disorders (SBD) | 3.70 (0.22) | 3.91 (0.33) | 3.55 (0.17) | 0.00 | 0.25 (-2 to 7) | 0.15 |
| Arousal Disorders (AD) | 3.18 (0.11) | 3.09 (0.07) | 3.15 (0.15) | 0.00 | 0.08 (-4 to 2) | 0.66 |
| Sleep-Wake Transition Disorders (SWTD) | 8.61 (0.45) | 8.15 (0.47) | 8.03 (0.44) | 0.00 | 0.03 (-5 to 5) | 0.84 |
| Excessive Daytime Sleepiness (EDS) | 6.58 (0.35) | 6.21 (0.38) | 6.15 (0.43) | 0.00 | 0.17 (-10 to 4) | 0.33 |
| Sleep Hyperhidrosis (HYH) | 3.85 (0.46) | 3.03 (0.42) | 3.30 (0.45) | 0.00 | 0.24 (-4 to 2) | 0.17 |
| Total | 37.67 (1.37) | 35.70 (1.38) | 34.88 (1.60) | 3.00 | 0.19 (-14 to 10) | 0.29 |
| **Behavior Rating Inventory of Executive Function (BRIEF) (n= 27 )** | | | | | | |
| Inhibit[b] | 49.07 (1.81) | 49.33 (2.06) | 50.19 (1.99) | 0.00 | 0.17 (-11 to 9) | 0.38 |
| Shift[b] | 46.63 (1.93) | 47.04 (2.38) | 47.22 (2.03) | -1.00 | 0.05 (-11 to 11) | 0.81 |
| Emotion control[b] | 41.04 (0.96) | 43.11 (1.31) | 41.70 (1.11) | 0.00 | 0.24 (-7 to 11) | 0.21 |
| Initiate[b] | 51.11 (1.89) | 49.67 (2.14) | 52.56 (2.17) | -2.00 | 0.36 (-24 to 12) | 0.06 |
| Working Memory[a] | 61.59 (1.63) | 57.44 (2.03) | 61.67 (1.68) | -4.22 (1.46) | 0.56 (-7.23 to -1.21) | 0.01 |
| Plan/Organization[a] | 58.44 (2.46) | 56.15 (2.63) | 59.15 (2.60) | -3.00 (1.64) | 0.35 (-6.37 to 0.37) | 0.08 |
| Organization of Materials[b] | 52.59 (2.09) | 49.67 (1.98) | 53.48 (2.16) | -7.00 | 0.42 (-25 to 10) | 0.03 |
| Self Monitor[a] | 56.78 (2.50) | 53.74 (2.89) | 58.07 (2.62) | -4.33 (1,97) | 0.42 (-8.38 to -0.28) | 0.04 |
| Behavioral Regulation | 44.81 | 45.93 | 45.63 | -2.00 | 0.01 (-7 to 10) | 0.97 |

| | | | | | | |
|---|---|---|---|---|---|---|
| Index (BRI)[b] | (1.39) | (1.90) | (1.49) | | | |
| Metacognition Index (MI)[a] | 57.81 (2.19) | 54.37 (2.54) | 58.78 (2.36) | -4.41 (1.62) | -0.53 (-7.73 to -1.08) | 0.01 |
| Global Executive Composite[a] | 52.78 (1.92) | 51.15 (2.34) | 53.85 (2.06) | -2.70 (1.29) | -0.41 (-5.35 to -0.06) | 0.05 |
| **Behavioral Inhibition /Activation System (BIS/BAS) (n= 31)** | | | | | | |
| BIS total[a] | 13.68 (0.66) | 13.32 (0.59) | 14.42 (0.65) | -1.10 (0.59) | -0.33 (-2.31 to 0.11) | 0.07 |
| BAS total[a] | 23.90 (1.04) | 25.23 (0.82) | 23.65 (0.83) | 1.58 (0.76) | 0.37 (0.03 to 3.13) | 0.05 |
| The fun-seeking subscale[b] | 7.35 (0.49) | 7,94 (0.51) | 7.35 (0.46) | 0.00 | 0.18 (-5 to 6) | 0.32 |
| The reward responsiveness subscale[b] | 8.61 (0.39) | 9.26 (0.37) | 8.39 (0.39) | 2.00 | 0.42 (-5 to 4) | 0.02 |
| The drive subscale[b] | 7.94 (0.48) | 8.03 (0.37) | 7.90 (0.44) | 0.00 | 0.13 (-5 to 4) | 0.46 |
